# Investigating the potential effect of antihypertensive medication on psychiatric disorders: a mendelian randomisation study

**DOI:** 10.1101/2020.03.19.20039412

**Authors:** Solal Chauquet, Michael O’Donovan, James Walters, Naomi Wray, Sonia Shah

**Author notes:** **Correspondance to:** Dr Sonia Shah, Institute for Molecular Biosciences, University of Queensland, Brisbane, QLD 4072, Australia.

## Abstract

**Background:** There is growing evidence from observational studies that drugs used for the prevention and treatment of CVD may cause, exacerbate, or relieve neuropsychiatric symptoms.

**Aim:** Use Mendelian randomisation (MR) analysis to investigate the potential effect of different antihypertensive drugs on schizophrenia, bipolar disorder and major depressive disorder.

**Methods:** We conduct two sample MR using expression quantitative trait loci (eQTLs) for antihypertensive drug target genes as genetic instruments, together with summary data from published genome-wide association studies, to investigate the causal effect of changes in drug target gene expression (as proxies of drug exposure) on psychiatric disorders.

**Results:** A 1 standard deviation lower expression of the *ACE* gene in blood was associated with 4.0 mmHg (95% CI = 2.7 – 5.3) lower systolic blood pressure, but increased risk of schizophrenia (OR (95% CI) = 1.75 (1.28 – 2.38)). A concordant direction of effect was observed with *ACE* expression in brain tissue.

**Conclusions:** Findings suggest an adverse effect of lower *ACE* expression on schizophrenia risk. This warrants further investigation to determine if lowering ACE activity for treatment of hypertension using ACE inhibitors (particularly centrally-acting drugs) may worsen symptoms in patients with schizophrenia, and whether there is any association between ACE inhibitor use and risk of (mainly late-onset) schizophrenia.

## INTRODUCTION

There is growing evidence for a bi-directional relationship between psychiatric disorders and cardiovascular disease (CVD). As a group, individuals with severe mental illness, including schizophrenia, bipolar disorder and major depressive disorder (MDD), have a 50% higher incidence of CVD^1^. On the other hand, depression is reported to be 3 times more common in patients after an acute myocardial infarction than in the general community^2^.

A higher cardiovascular morbidity and mortality observed in individuals with psychiatric disorders has been partly attributed to both a higher incidence of CVD risk factors, and less effective clinical management^3^. For example, the Recovery After an Initial Schizophrenia Episode (RAISE) study of 394 patients with first-episode schizophrenia spectrum disorders reported that 48% were obese or overweight, 56% had dyslipidemia, 40% had prehypertension and 10% had hypertension^4^. There is therefore great need to treat these patients for CVD prevention. The high prevalence of CVD risk factors may be due to unfavourable effects of anti-psychotic medication, adverse dietary habits, lack of exercise and increased substance use (a meta-analysis of 42 studies reported a smoking prevalence of 62% in schizophrenia^5^). However, there is also evidence of shared pathophysiological mechanisms. A genome-wide association study (GWAS) of bipolar disorder, for example, identified genetic risk variants within the *CACNA1C* gene, whose encoded protein is also the target of calcium channel blockers used for the treatment of hypertension^6,7^. The directionality of effect, if any, of this class of drugs on bipolar disorder remains unclear^8,9^. Observational studies have reported associations between antihypertensive medication and psychiatric disorders, though the reported direction of association appears to be drug-class dependent^10,11^, suggesting that any effect could be independent of their blood pressure lowering effect.

The gold standard approach for determining a causal effect of drug treatment would be randomised control trials (RCTs). However, RCTs are expensive to conduct and may not be feasible for various reasons, which may explain a current lack of large, high-quality RCTs investigating the effects of antihypertensive medication on symptoms in patients with psychiatric disorders or in relation to disease risk itself. Mendelian randomisation (MR) is a form of instrumental variable analysis that uses genetic variants that are robustly associated with a potentially modifiable exposure as unconfounded instruments to distinguish whether an observed association between the exposure (e.g. drug treatment) and outcome (e.g. disease) is causal or not. Given all assumptions are satisfied, as discussed below, MR analysis can be used to overcome two major limitations of evidence from observational studies: unmeasured confounding and the ability to distinguish cause from consequence^12^.

Assuming that lower mRNA levels of a gene translates into lower protein levels and subsequently lower protein activity, a single genetic variant associated with gene expression (expression quantitative trait locus (eQTL)) of a drug target gene can be used as an instrument for drug exposure^13^. For example, a sample population can be split into two groups - those that carry the allele associated with lower gene expression (equivalent to the treatment arm of an RCT where the drug is an inhibitor), and those that do not carry any expression-lowering alleles (control arm). Just as would be done in an RCT, incidence of disease over time in these two groups can then be compared (**Figure 1**). The utility of a single nucleotide polymorphism (SNP) as an instrument for drug treatment has recently been demonstrated using lipidomics data. In that proof-of-concept study, a SNP in the *HMGCR* gene, used as a genetic instrument for HMGCR inhibition by cholesterol-lowering statin medication, gave rise to a strikingly similar association pattern in a healthy sample, to actual changes in plasma lipoprotein subclasses and lipid fractions in an independent sample before and after statin therapy^14^. The same genetic variant was also used to investigate the potential effect of statin treatment on the risk of type 2 diabetes^15^. In addition to single SNPs as instruments, the MR framework can be extended to use multiple independent SNPs associated with polygenic exposures, for example to investigate the effect of systolic blood pressure on valvular heart disease^16^.

**Figure 1.**
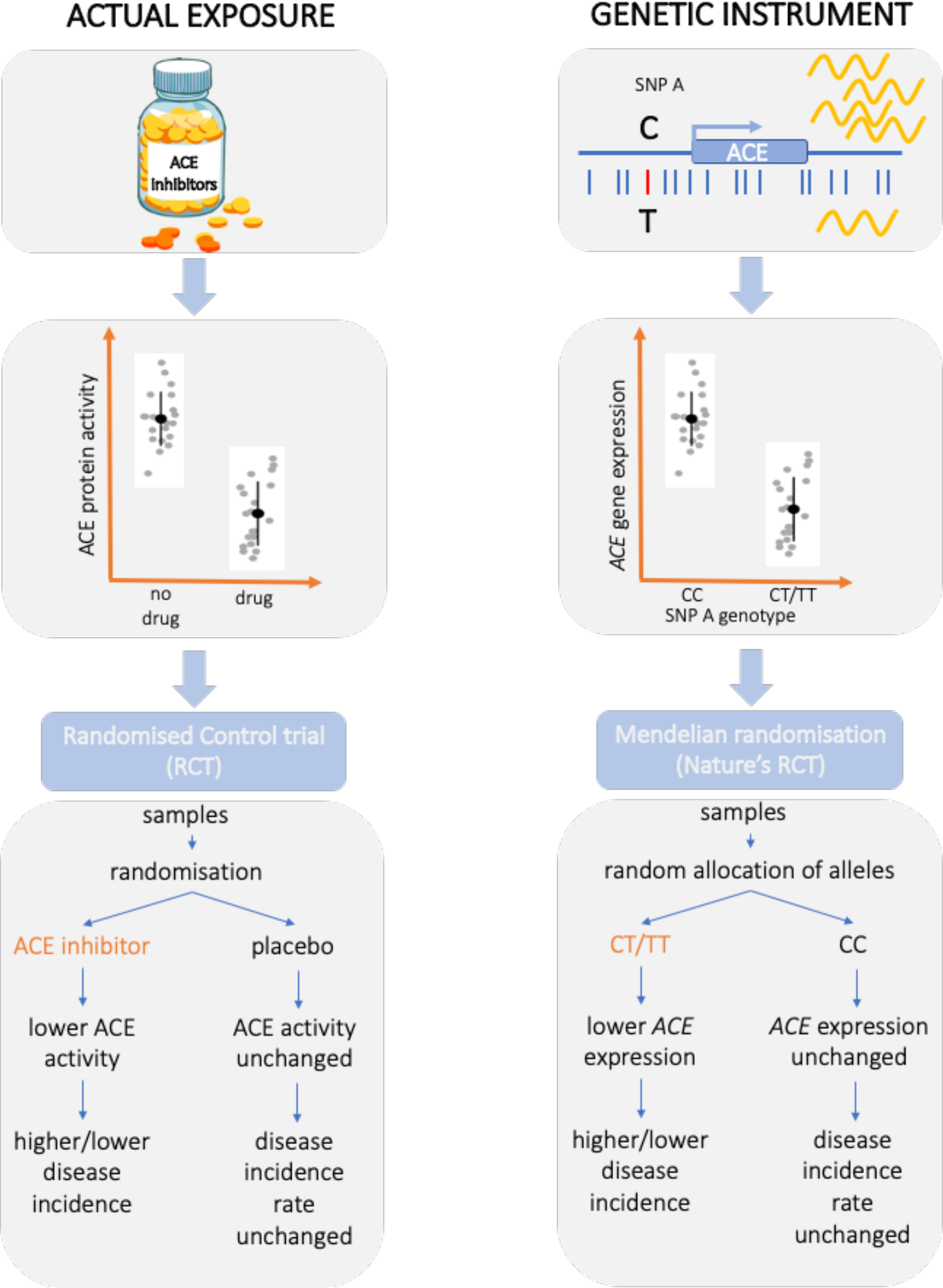
Mendelian randomisation vs randomised control trial. The expected outcome from a hypothetical randomised control trial (RCT) of ACE inhibitors (left) compared to Mendelian randomisation analysis using a single nucleotide polymorphism (SNP) associated with *ACE* gene expression (expression quantitative trait loci (eQTL)) as an instrument for exposure to ACE inhibitors (right). The eQTL SNP may exert effects on gene expression by affecting binding of transcription regulators. The presence of the T allele is associated with lower expression of *ACE*. Assuming that the difference in gene expression translates to a difference in protein activity, individuals that carry a T allele at this eQTL SNP (genotypes ‘CT’ or ‘TT’) are therefore comparable to the treatment arm of the RCT study, while those that do not carry any T allele (genotype ‘CC’) to the control arm.

MR analysis makes three major assumptions that need to be met. Firstly, the instrument is robustly associated with the exposure of interest. Weak instruments can lead to bias, and selection of instruments should assess instrument strength. Second, there are no unmeasured confounders between genetic instruments and outcome. Sufficient adjustment for population stratification in GWAS can help minimise confounding from ancestry, while identifying association of instruments with other covariates may help to identify and assess other potential sources for confounding. Finally, the instrument should not affect the outcome except through the biomarker of interest. The assumptions of an MR analysis hold if the associations of a genetic variant with other covariates arise because of the serial and sequential effects of the gene of interest on others residing more distally on the same causal pathway to disease^17^ (i.e. association through vertical pleiotropy rather than horizontal pleiotropy). It is plausible for a single genetic variant to meet these conditions if the biological process linking the variant with the exposure is well understood^12^. Where mRNA or protein levels are the primary exposure of interest, any pleiotropy observed of a *cis*-regulatory SNP instrumenting its encoded mRNA or protein is more likely to be vertical than horizontal in origin^17^.

MR analysis requires an estimation of the effect of the genetic instrument on exposure as well as on outcome. Traditionally, this required genetic information, exposure and outcome all measured in the same individuals (single sample MR), which is not always feasible, especially to achieve the large sample sizes needed for well-powered studies. Two-sample MR uses two independent samples, one to estimate the effect of the genetic instrument on exposure and the other on outcome. This approach enables us to use summary results from GWAS based on very large sample size which produce more precise estimates.

Given the high prevalence of hypertension in psychiatric disorders and emerging evidence for a role of some antihypertensive drug target genes in the aetiology of specific disorders, understanding whether these medications may cause, exacerbate, or relieve neuropsychiatric symptoms^18^ will enable clinicians to make better-informed prescription decisions in high-risk individuals. In this study, we use publicly available eQTL data to identify suitable genetic instruments for expression of blood pressure (BP)-lowering drug target genes. We combine this with summary data from publicly available GWAS on psychiatric disorders, in a two-sample MR analysis to investigate the effect of changes in drug target gene expression (as proxies for exposure to antihypertensive drugs) on psychiatric disorders risk.

## METHODS

### Identification of drug target genes

Using the WHO Collaborating Centre (WHOCC) for Drug Statistics Methodology we identified the different classes of BP-lowering drugs, as classified by the Anatomical Therapeutic Chemical (ATC) classification system. Using the same website, we obtained a list of active ingredients within each drug class. Genes whose protein products are targeted by any one of these active ingredients were identified using the DrugBank^19^ (https://www.drugbank.ca/) and ChEMBL^20^ (https://www.ebi.ac.uk/chembl/) databases. If a gene was named as a target in only one database, we looked at the referenced publications to determine if experimental evidence was sufficient for inclusion as a drug target gene.

### Genetic instruments for drug target gene expression (exposure)

We used publicly available eQTL data (**Supplementary Table 1**) to identify single nucleotide polymorphisms (SNPs) associated with expression of antihypertensive drug target genes. Though functional analysis of GWAS summary data for psychiatric disorders suggest brain tissues to be the most relevant for disease aetiology^21,22^, eQTL datasets based on expression in brain tissues have only modest sample numbers and therefore less power to identify robust SNP instruments for use in MR studies. The GTEx project^23^ is a publicly available resource for eQTL data generated from multiple different tissues, with sample sizes ranging from around 139-255 for individual brain regions, while the PsychENCODE resource provides eQTLs measured in ∼1300 prefrontal cortex samples (**Supplementary Table 1**). In comparison, the eQTLGen consortium has generated the largest eQTL dataset to date from blood samples (N = 31,684)^24^. A study comparing eQTLs between blood and brain found fairly high concordance between the two datasets, providing rationale for the use of blood eQTL data and its suitability for making inferences for brain-related diseases^25^. In other words, although genes may have enhanced expression in brain compared to blood, many genes show some expression in blood, and SNPs that control variation of gene expression between people in brain tissues are likely to also control inter-individual variation in gene expression in blood. Hence, for each of the drug target genes we queried the eQTLGen dataset to identify the most significant SNP instruments that were associated with the expression of these genes in blood and followed up statistically significant associations using the same approach using eQTL data from brain tissues (see below). Only *cis-*associations are available in the eQTLGen data (associations where the distance between SNP and gene is <1 megabase (Mb)). The eQTL data is on the scale of a standard deviation (SD) change in the expression level of the gene for each additional effect allele.

### Assessing Instrument strength

The F-statistic from the regression of the exposure (gene expression) on the instrument (eQTL SNP) is usually quoted in single sample MR studies as a measure of the strength of an instrument. By rule of thumb, instruments with an F-statistic less than 10 are considered ‘weak instruments’^26^. A value of 10 indicates that bias in the estimated causal effect due to measurement error is around 10% of the true value of causal effect. In the context of two-sample MR which uses GWAS summary data, the F-statistic can be generated using the approximation described by Bowden et al^27^. F-statistic for SNP *j* can be approximated as 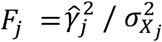 where 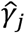 is the SNP-exposure association and 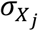 is the standard error of the SNP-exposure association.

### Outcome data

We obtained publicly available case-control GWAS summary data for schizophrenia (40,675 cases, 64,643 controls)^21^, bipolar disorder (20,352 cases, 31,358 controls)^28^ and MDD (135,458 cases, 344,901 controls)^22^ (**Supplementary Table 2**). Further details of samples and GWA analyses are provided elsewhere ^21,22,28^.

### Two-sample MR analysis

To perform two-sample MR analysis between exposure (gene expression) and outcome, we used the Summary-based mendelian randomisation (SMR) method^29^. Briefly, let *y* be the outcome of interest, *x* be gene expression (exposure), *g* be a genetic instrument, *b*_*gx*_ be the effect of *g* on *x*, and *b*_*gy*_ be the effect of *g* on *y*. In a single sample MR analysis, the effect of exposure *x* on outcome *y* free of any non-genetic confounders, is defined as *b*_*gy*_/*b*_*gx*_. The latter can also be estimated and tested using GWAS summary-level data in a two-sample MR framework, as is done by the SMR method. The SMR approach selects a single *cis*-eQTL SNP that is most significantly associated with gene expression as an instrument. All analyses were performed using SMR version 1.02.

### Assessing association due to linkage

In the absence of confounding factors such as population stratification and assortative mating, MR can estimate an apparent effect of an exposure on the outcome in 3 scenarios: vertical pleiotropy (causality); horizontal pleiotropy, where the same SNP influences the exposure and outcome through independent pathways (e.g. if the same SNP affects expression of two different genes, but only one gene is associated with outcome); or linkage, where the SNP that influences the exposure is in linkage disequilibrium (LD) with another SNP that independently influences the outcome. We used the heterogeneity in dependent instruments (HEIDI) test implemented in the SMR tool to identify association between gene expression and outcome due to a linkage scenario^29^. Briefly, if gene expression and a trait share the same causal variant, the *b*_*xy*_ values calculated for any SNPs in LD (using the default value of *r*^*2*^ > 0.05 and also *r*^*2*^ < 0.9 to avoid issues of collinearity) with the causal variant should be identical. Therefore, testing against this null hypothesis of a single causal variant is equivalent to testing for heterogeneity in the *b*_*xy*_ values estimated for the SNPs in the *cis*-eQTL region^29^. Since heterogeneity estimates may not be robust if using only a small number of SNPs, the HEIDI test required a minimum of 5 SNPs for estimating heterogeneity. As recommended by the authors, a HEIDI test p-value < 0.01 was considered to indicate heterogeneity in *b*_*xy*_ values, suggesting that association between gene expression and outcome is most likely to be due to a linkage scenario. SMR requires a reference dataset from which to estimate LD between SNPs. For this purpose, a random sample of 10,000 unrelated individuals of European ancestry from the UK Biobank data was used.

### Genetic instrument validation

Using eQTL data as a proxy for drug exposure assumes that changes in gene expression translate into changes in protein activity and subsequently the intended outcome of drug exposure. This may not necessarily be the case as post-transcriptional mechanisms may affect function independent of transcriptional changes. Therefore, to validate our genetic instruments and show that changes in gene expression reflect the effect of drug exposure, we performed SMR analysis with blood gene expression as exposure and systolic blood pressure (SBP) as the outcome using summary results from a GWAS in 757,601 individuals of European ancestry^30^ (**Supplementary Table 2**). Genes whose expression in blood was not associated with SBP with at least nominal significance (i.e. SMR p-value ≥ 0.05) were excluded from further analysis. The effect estimate from SMR of gene expression on SBP represents mmHg change in SBP per 1 SD increase in gene expression.

### MR analysis with blood gene expression and psychiatric outcomes

We ran SMR analysis to estimate the effect of a SD change in drug target gene expression levels in blood (using the eQTLGen data) on outcomes of interest (using GWAS summary data for schizophrenia, bipolar disorder and MDD). Main results are presented as the effect on disease risk (odds ratio) per 1 SD change in gene expression, where the direction of gene expression change is harmonised to reflect an SBP-lowering effect. The effect estimates in the main results therefore show the potential direction of effect of antihypertensive drug exposure on risk of psychiatric disorders. Bonferroni correction for multiple testing was used to identify significant associations.

### Sensitivity analyses

#### Colocalisation analysis

For any statistically significant MR associations, we applied a Bayesian colocalisation approach to estimate the posterior probability for a common causal variant for gene expression and outcome^31^ using the coloc (v3.1) R package (https://cran.r-project.org/web/packages/coloc/). Since for each gene, SNP associations in blood were only available for those within a 2MB window, colocalisation analysis was restricted to using only these *cis*-SNP associations. Default priors were used for analysis.

#### Assessing horizontal pleiotropy

The same genetic variant could be associated with expression of more than one gene, which could invalidate the assumption that the genetic instrument is associated with outcome only via changes in gene expression of the drug target gene. To assess the presence of horizontal pleiotropy, for each genetic instrument we also extracted available associations with all other nearby genes (within a 2Mb window). For genes whose expression was associated at nominal significance (p < 0.05), we performed SMR analysis to determine if the expression of these genes was associated with the outcome of interest, and colocalisation analysis to determine the posterior probability of a shared causal variant.

#### MR analysis using a proxy for the ACE insertion/deletion as an instrument

Previous studies have looked at the association of the ACE insertion/deletion (I/D) in relation to psychiatric disorders. The ACE indel consists of the presence or absence of a 250-bp DNA fragment in intron 16. The presence of D allele is associated with higher ACE enzyme activity in comparison to I allele, accounting for almost 50% of the variation in plasma ACE levels^32^. The SNP rs4343 is considered the best proxy for the *ACE I/D*^33^, with the A allele corresponding to the insertion and the G allele to the deletion. The G allele of the rs4343 SNP has been associated with increased plasma ACE activity accounting for 16.2% of the total variance in ACE activity^34^, as well as increased ACE protein levels in cerebral spinal fluid^35^. Linkage disequilibrium (LD) between rs4343 and the best eQTL SNP selected by the SMR method as an instrument for *ACE* expression in blood (rs4277405), is around *r*^*2*^ 0.35 in Europeans (LD estimation from LDlink https://ldlink.nci.nih.gov/). We performed MR analysis using rs4343 as an instrument for *ACE* expression in blood to determine if consistent effects were observed on schizophrenia risk.

#### Analysis of ACE protein levels with gene expression and schizophrenia risk

To determine if genetic variants associated with *ACE* mRNA expression in blood also associated with ACE protein levels we performed SMR and co-localisation using eQTL association in blood (eQTLGen data) and published GWAS summary data of ACE protein levels in cerebral spinal fluid (CSF) (Supplementary Table 2). We then performed SMR and colocalisation analysis between genetic variants associated with ACE protein levels in CSF and schizophrenia risk. All analyses were restricted to SNPs within a 2Mb window around the *ACE* gene due to availability of only *cis*-associations in the eQTL data.

#### Assessing confounding due to ancestry

One potential confounder in MR analyses is ancestry. For both the bipolar disorder and MDD GWAS analysis was restricted to individuals with European ancestry. For schizophrenia, though a small proportion of individuals consisted of non-European ancestry, analysis was performed using using matched ancestry controls, and within-ancestry analysis prior to meta-analysis to avoid confounding. As sensitivity analyses we ran SMR analysis using ACE eQTL data and case-control GWAS summary data generated from a subset of individuals of European ancestry only (33,640 schizophrenia cases and 43,456 controls) and data from East Asian ancestry individuals only (22,778 schizophrenia cases and 35,362 controls) ^36^ (**Supplementary Table 2**). A random sample of 10,000 unrelated individuals of European ancestry and all unrelated East Asian samples from the UK Biobank were used as an LD reference datasets for SMR analysis.

### MR analysis with brain gene expression and psychiatric outcomes

Given that expression in brain is most relevant for psychiatric disease, for any statistically significant associations observed with blood eQTL data, we also performed MR analysis using expression data in brain tissue. We used eQTL data from the PsychENCODE resource (**Supplementary Table 1**), which is a meta-analysis of eQTL studies of brain prefrontal cortex samples, with a total sample size of 1,387^37^. Only associations for SNPs located within a 1 Mb window around each gene are available in the PsychENCODE data. Given different brain regions may have greater relevance to different diseases, we also ran SMR analysis using gene expression in 13 different brain regions from the latest version of GTEx (v8), with sample size ranging between ∼130 - 250. For GTEx, data is only available for SNP-gene associations within a 2 Mb window around the transcription start site (**Supplementary Table 1)**.

### MR analysis of other genes in the renin-angiotensin pathway

*ACE* is part of the renin-angiotensin (RAS) pathway that regulates blood pressure. To determine whether other components of the pathway show any association with schizophrenia, we performed MR analysis on other genes in the RAS pathway (*AGT, REN, AGTR1*), regardless of whether they are targets of antihypertensive medication or not. *AGT, AGTR1* and *REN* were absent in the blood eQTL summary data. All 3 genes were present either in the PsychENCODE or GTEx brain data. We therefore identified the strongest eQTL SNPs for these genes in the brain eQTL datasets and performed SMR analysis with schizophrenia as an outcome.

### MR analysis of systolic blood pressure as exposure and schizophrenia risk

To determine whether the effect of *ACE* on risk of schizophrenia is likely to be mediated via changes in blood pressure or whether the effect may be independent of blood pressure, we estimated the effect of SBP on schizophrenia using summary data from the largest available GWAS in European individuals on SBP^30^ (**Supplementary Table 2**) and the Generalised Summary-data-based MR (GSMR) method^38^. GSMR is an extension of the SMR method that uses multiple genetic variants associated with the risk factor to test for putative causality. Lack of evidence of an MR association would suggest that any effect of ACE on schizophrenia is likely to be independent of its effect on blood pressure.

### Patient and Public Involvement

As this analysis used only published summary data from studies involving human participants (that provided written informed consent and published studies were approved by their respective institutional ethics review committees), it was not appropriate or possible to involve patients or the public in the design, or conduct, or reporting, or dissemination plans of our research.

### Data availability

All datasets used in this analysis are publicly available and sources for each are provided in supplementary tables.

## RESULTS

### Genetic instrument selection

We identified a total of 110 genes whose encoded protein activity has been experimentally shown to be modified by one or more BP-lowering drugs (**Supplementary Table 3**). Though 109 out of 110 were reported by eQTLGen to be expressed in blood, 48 of the protein encoding genes were absent in the meta-analysed eQTLGen summary data as they showed no variation in expression across samples and were filtered out during meta-analysis. We were therefore only able to query the eQTL data for 61 out of the 110 drug target genes. For all 61 genes, the most significant *cis-*eQTL SNPs selected as genetic instruments had an F-statistic > 10 (**Supplementary Table 4**). Out of the 61 genes, 22 had gene expression levels in blood that were associated with SBP at nominal significance (SMR p-value < 0.05) (**Supplementary Table 4**), and these were taken forward to the main analysis.

### MR analysis with blood gene expression and psychiatric outcomes

Using a significance threshold of p-value < 7.6 × 10^−4^ (Bonferroni correction for association testing of 22 genes with 3 disease phenotypes) and a HEIDI p-value ≥ 0.01 to rule out association due to linkage, we found a 1 SD decrease in blood *ACE* expression (target for ACE inhibitors) was associated with 4.0 mmHg (95% CI = 2.7 – 5.3) lower SBP and a higher risk of schizophrenia (OR (95% CI) = 1.75 (1.28 – 2.38), p-value = 3.95×10^−4^) (**Figure 2**). Colocalisation analysis gave a posterior probability of 85% for a common causal variant for *ACE* expression and schizophrenia risk. Though expression of *CYP17A1* (targeted by Spironolactone, a potassium-sparing diuretic) and *NISCH* (targeted by Moxonidine, a new-generation alpha-2/imidazoline receptor agonist) was associated with both schizophrenia and bipolar disorder, the significant HEIDI p-values suggest these associations are likely due to linkage. This is corroborated by the low to moderate posterior probabilities for a common causal variant for these associations (<0.1% for *CYP17A1* expression and schizophrenia risk, 53% for *CYP17A1* expression and bipolar disorder risk, <0.1% for *NISCH* expression and schizophrenia risk, 36% for *NISCH* expression and bipolar disorder risk). These associations were therefore not taken forward for further analyses. SMR results from the MR analysis between expression of the 22 drug target genes in blood with the three psychiatric disorders are provided in **Supplementary tables 5 – 7**.

**Figure 2.**
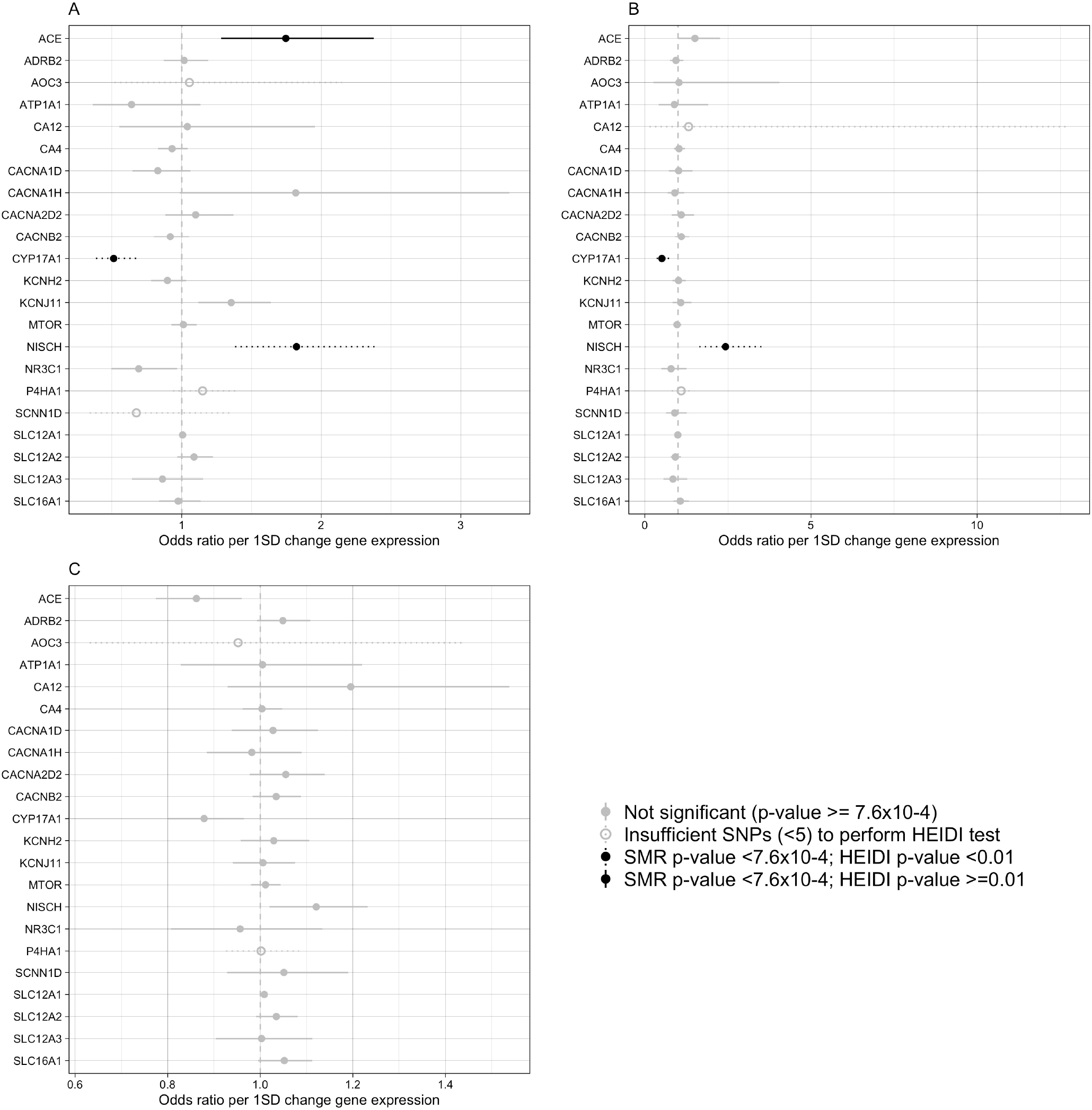
Association of drug target gene expression in blood with disease risk. Forest plot of the effect of a standard deviation (SD) change in expression of 22 BP-lowering drug target genes in blood on A) schizophrenia B) bipolar disorder C) major depressive disorder risk (odds ratio). Bars represent 95% confidence intervals. The direction of gene expression change reflects the SBP-lowering effect. An odds ratio > 1 suggests an increased risk of disease associated with the direction of gene expression change that is also associated lowering of blood pressure. Associations that are statistically significant after correcting for multiple testing (22 genes × 3 phenotypes; p-value < 7.6 × 10^−4^) and that have a HEIDI p-value >= 0.01 are shown by black lines. Associations that are statistically significant after correcting for multiple testing but have a HEIDI p-value < 0.01 (indicating association likely due to linkage) are shown in black dotted lines. Associations that did not pass the multiple testing correction are shown in grey lines. Associations for which a HEIDI test could not be performed due to unavailability of sufficient number SNPs (<5) are shown by grey dashed line.

### Assessing horizontal pleiotropy

The eQTL SNP instrument for *ACE* expression in blood (rs4277405) was associated with the expression of several adjacent genes (*FTSJ3, TANC2, SMARDCD2, ICAM2* and *STRADA*) in blood (**Supplementary Table 8**). However, SMR analysis did not provide any evidence for association between expression of these adjacent genes with schizophrenia (**Supplementary Table 9**). Only *ACE* expression was significantly associated with schizophrenia and also had the highest posterior probability for a shared causal variant - a posterior probability of 85% for *ACE* expression and schizophrenia compared to a posterior probability < 5% for all other nearby genes (**Supplementary Table 9**). The localisation of the *ACE* eQTL signal and GWAS signal is illustrated in **Supplementary Figure 1**.

### MR analysis using a proxy for the ACE insertion/deletion as an instrument

The G allele of rs4343 (proxy for the ACE deletion) has previously been reported to be associated with higher ACE protein levels in CSF^35^ and higher plasma ACE enzyme activity^34^. In blood eQTL data, the G allele was associated with higher *ACE* gene expression levels, showing concordant direction of effect of the G allele from gene expression to protein levels to activity in independent datasets. Using rs4343 as an instrument, we observed a concordant direction of effect of 1 SD lower *ACE* gene expression in blood on lower SBP (beta (SE) = -2.46 mmHg (0.79); p-value = 1.7 × 10^−03^), but higher risk of schizophrenia (OR (95% CI) = 1.84 (1.16 – 2.91), p-value = 9.4×10^−03^) (**Supplementary Table 10**).

### Analysis of ACE protein levels with gene expression and schizophrenia risk

A 1 SD increase in *ACE* blood expression was associated with a 4.99 ng/ml increase in ACE protein levels in CSF (p-value = 3.6 ×10^−07^; HEIDI p-value = 0.42), and the posterior probability for a shared causal variant was 99.3%. Lower ACE CSF protein levels were associated with higher risk of schizophrenia (OR per 1 ng/ml (95% CI) = 1.12 (1.05 – 1.19); p-value = 9.4 ×10^−04^; HEIDI p-value = 0.39), with a posterior probability for a shared causal variant of 80.1% (**Supplementary Table 11**).

### MR analysis using schizophrenia GWAS in European and East Asian ancestry individuals

A 1 SD decrease in blood *ACE* expression was associated with increased risk of schizophrenia using GWAS summary data from both European ancestry individuals (OR (95% CI) = 1.78 (1.25 – 2.51); p-value = 1.2 × 10^−03^) and East Asian ancestry individuals (OR (95% CI) = 2.69 (1.62 – 4.48); p-value = 1.3 × 10^−04^) (**Supplementary Table 12**).

### MR analysis of ACE brain expression and psychiatric outcomes

The eQTL SNP instrument for expression of *ACE* in brain prefrontal cortex (PsychENCODE data) had an F-statistic of 43.9 (**Supplementary Table 13**). Lower *ACE* expression in prefrontal cortex was associated with higher risk of schizophrenia (OR (95% CI) = 1.33 (1.13 – 1.56), p-value = 5.×10^−04^), concordant with that observed in blood (**Figure 3**). Extending analysis to other brain regions, significant associations after multiple testing between *ACE* expression with schizophrenia risk were observed for cortex regions, cerebellum, and cerebral hemisphere (**Figure 3**). A 1 SD decrease in prefrontal cortex *ACE* expression was associated with increased risk of schizophrenia in both European ancestry (OR (95% CI) = 1.34 (1.12 – 1.60); p-value = 1.5 × 10^−03^) and East Asian ancestry individuals (OR (95% CI) = 1.59 (1.22 – 2.07); p-value = 6.0 × 10^−04^) (**Supplementary Table 12**).

**Figure 3.**
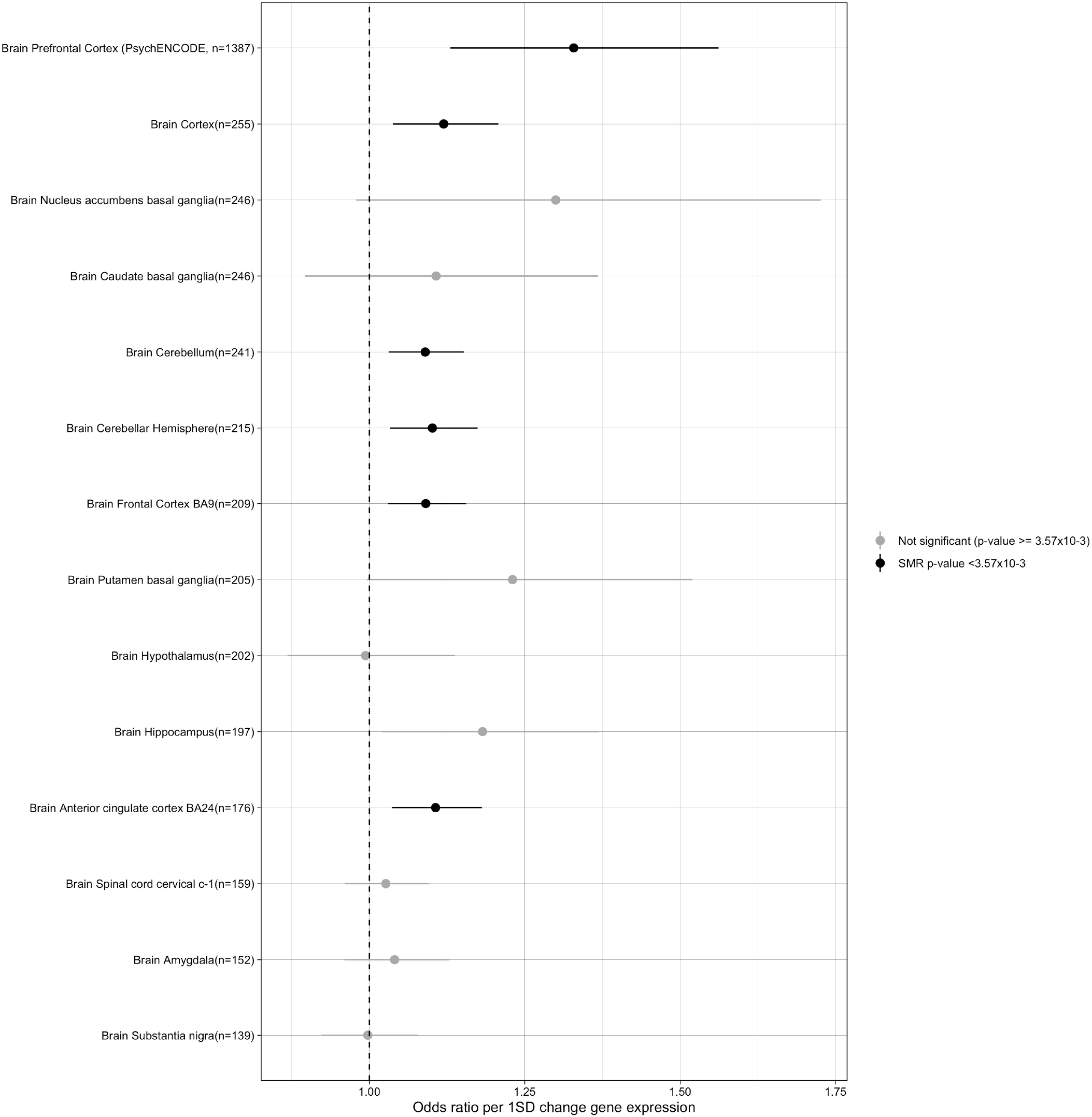
MR analysis of *ACE* expression in brain tissue with risk of schizophrenia. Forest plot of the effect of a 1 standard deviation (SD) decrease in *ACE* gene expression on schizophrenia risk in different brain regions. eQTL data was queried for 13 different brain regions from GTEX v8 and the prefrontal cortex region from PsychENCODE. Associations that are statistically significant after correcting for 14 tests (p-value < 3.57×10^−3^) are shown in black. The sample size (n) for each dataset is provided in brackets.

### MR analysis of other genes in the renin-angiotensin pathway and schizophrenia risk

Since *AGT, AGTR1* and *REN* were absent in the blood eQTL summary dataset, we queried brain eQTL data. eQTL SNP instruments for expression of *AGT, AGTR1* and *REN* in brain tissue all had F-statistics > 10, though these varied from 13.4 for *REN* to 469.6 for *AGT*. There was lack of evidence for association between expression of these three RAS pathway genes in brain tissue and schizophrenia risk (**Supplementary Table 12**).

### MR analysis of blood pressure and schizophrenia risk

Using >800 SNPs that are independently associated at genome-wide significance (GWAS association p-value < 5×10^−8^) with SBP as MR instruments, we found little evidence to support a causal effect of SBP on schizophrenia (OR (95% CI) = 0.998 (0.996 – 1.00) ; p-value = 0.14)) (**Supplementary Figure 2**).

## DISCUSSION

Mendelian randomisation uses genetic variation to investigate causal relations between potentially modifiable exposures and health outcomes. In this study we use published summary data from eQTL and GWA studies in a two-sample MR analysis to make inferences about the potential effects of antihypertensive medication, widely used for the treatment and prevention of CVD, on psychiatric disorders. Lower expression of the *ACE* gene was associated with decreased systolic blood pressure but increased risk of schizophrenia. A summary of the study design and results is shown in **figure 4**. There was lack of evidence to support a causal effect of blood pressure itself on schizophrenia risk, suggesting that any effect of *ACE* on schizophrenia risk may be independent of blood pressure regulation. Concordant effects of *ACE* expression in brain tissue on schizophrenia risk were observed, with strongest association in cortex and cerebellum regions. While the prefrontal cortex has consistently been implicated in the pathophysiology of schizophrenia, recent evidence also suggests a role for the cerebellum in higher order cognitive functions as well as psychopathology^39,40^. We observed concordant effects in two independent samples of European-only and East Asian-only ancestry.

**Figure 4.**
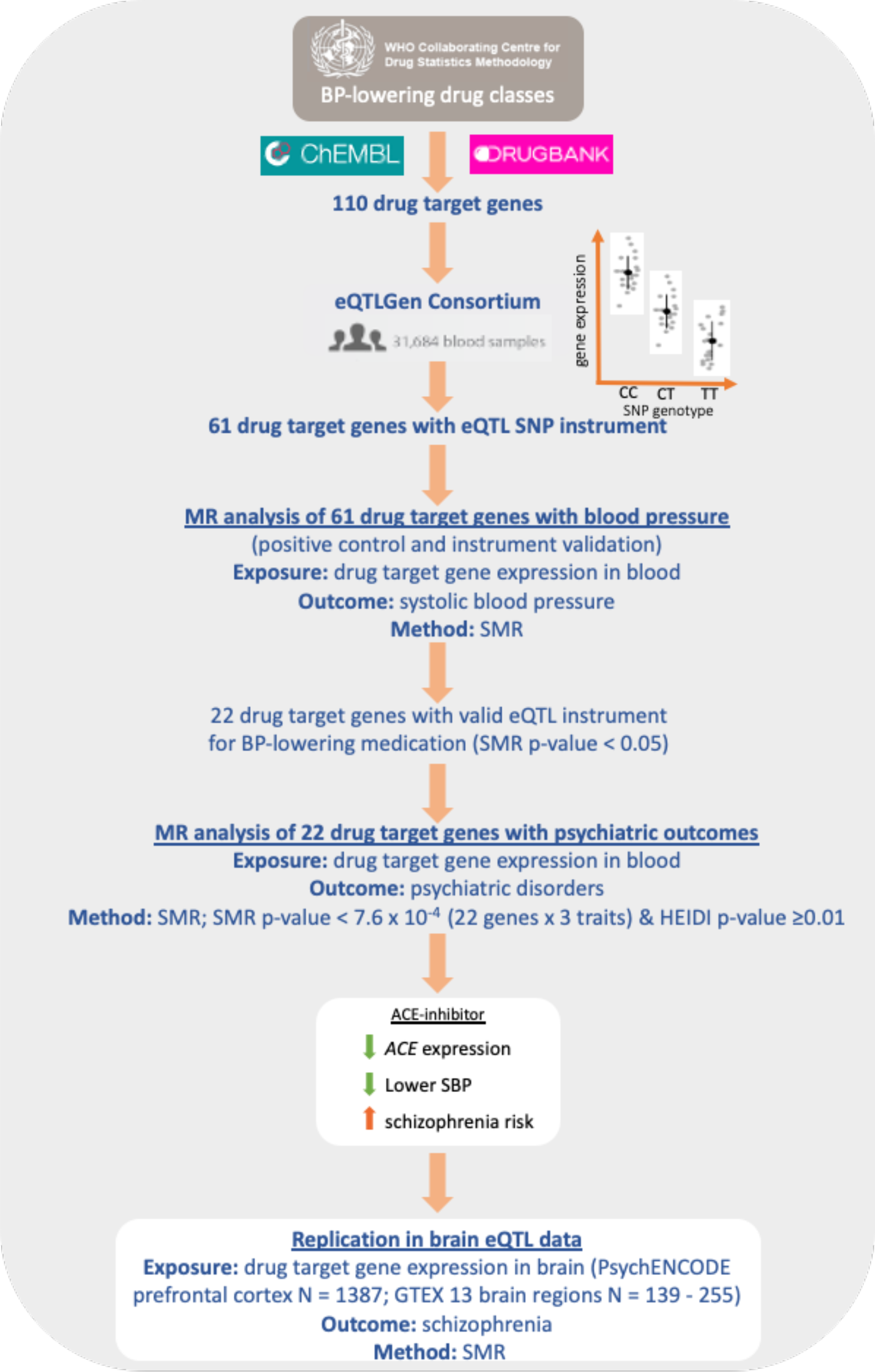
Summary of study design and results.

*ACE* is a component of the renin-angiotensin system (RAS) that regulates blood pressure. In addition to the ‘classical’ circulating RAS, nearly all organs of the body have their own local paracrine-like RAS, with organ-specific actions, including the brain^41^. Though we were unable to test the association between expression of other RAS components in blood, there was lack of evidence for association between expression in brain tissues of other RAS pathway genes, namely *AGT, AGTR1* and *REN*, and schizophrenia risk. However, given the small sample numbers and varying F-statistics, a role of some RAS components (namely AGTR1 and *REN*) cannot be ruled out, and larger brain eQTL datasets may be required to confirm small, but clinically relevant effects. ACE consists of two catalytically independent domains (N-domain and C-domain) with high sequence similarity. In addition to the cleavage of angiotensin I to angiotensin II, the two domains have been shown to bind multiple substrates with varying affinities^42^. The location of ACE on the cellular membrane of neurons and its ability to cleave multiple different substrates, such as bradykinin (BK), neurotensin (NT), substance P, enkephalins (EKs), amyloid beta and other hormone/neuropeptides, suggests a role for ACE in central nervous system functions^43^. ACE has been shown to play a role in both innate and adaptive immune responses by modulating macrophage and neutrophil function^44^, which may have relevance given the increased evidence of the role of inflammation and immunity in schizophrenia^45^. *In vitro* studies have shown elevated amyloid-beta protein levels following ACE inhibition^46^, raising questions on the safety of ACE-inhibitors in Alzheimer’s disease patients or those with a family history of the disease. Our findings raise a similar concern in relation to schizophrenia.

### Comparison with other studies

An MR analysis of antihypertensive drug exposure and risk of psychiatric disorders has not been previously conducted. Large, high-quality RCTs on this are also lacking, most likely due to feasibility issues. Only a few observational studies have investigated the effects of antihypertensive therapy on risk of psychiatric disorders. Boal et al looked at four drug classes in relation to mood disorders and found that calcium antagonists and beta-blockers were associated with increased risk of mood disorder admissions^10^. Though they also looked at ACE inhibitors, users were grouped together with ARB users, and together this group had the lowest risk for mood disorder admissions. Hayes et al^11^ found evidence that exposure to L-type calcium channel antagonists reduced rates of psychiatric hospitalization and self-harm in patients with bipolar disorder and schizophrenia, but did not look at other antihypertensive drug classes. On the contrary, a systematic review looking at data from 23 studies, with six randomised, double-blind, controlled clinical trials, all of which investigated verapamil, a calcium channel blocker, in acute mania, found no evidence of an effect^8^.

Previous genetic studies have identified an association between the *ACE* insertion/deletion (*I/D*) and schizophrenia, however the findings have been contradictory. Cresenti et al^47^ and Mazaheri et al^48^ found the *D* allele to be protective in a Spanish and Iranian population, respectively. Kucukali et al^49^ found the *I* allele to be protective in a Turkish sample, while Song & Lee^50^ found little evidence of an association based on a multi-ethnic meta-analysis of 13 studies. Similarly, paradoxical findings can be found for ACE activity and protein levels, as both reductions and increases have been reported when comparing patients with schizophrenia with controls^51–55^. Given the small sample size and observational nature of these studies, they are not able to distinguish whether lower ACE expression or activity plays a causal role or whether the observed differences are simply a consequence of disease, anti-psychotic treatment or other unmeasured confounders. Our MR analysis suggests a possible role for *ACE* in disease aetiology that is independent of its effect on blood pressure. Though the most recent GWAS for schizophrenia^21^ did not identify any variants around the *ACE* gene at genome-wide significance (p-value < 5×10^−8^), there was suggestive evidence for genetic association at this locus (most strongly associated genetic variant with an association p-value of 5.46×10^−5^), and larger sample sizes would be needed to identify a robust genetic association.

### Strengths of this study

Two sample MR analysis allows us to use the largest available GWAS and eQTL datasets to investigate causality by overcoming some of the caveats of observational studies and RCTs, such as sample size, confounding bias and feasibility. As mentioned previously, MR analysis requires three assumptions to be met. The first assumption requires the instrument to be associated with exposure. We assessed instrument strength using the F-statistic as well as selecting instruments that show an intended effect on blood pressure itself. The second assumption is that confounders are not associated with the instrument. Population stratification is one such confounder. We believe that eQTL studies and GWAS studies of psychiatric disorders have sufficiently corrected for population stratification. Both the GWAS on bipolar disorder and major depressive disorder have been restricted to European ancestry and adjusted for principal components. The schizophrenia GWAS included predominantly individuals with European ancestry, however a small proportion of individuals also have East Asian and African American ancestry. We believe the published meta-analysis was carried out carefully to avoid confounding. As sensitivity analyses we performed MR analysis on summary data generated from meta-analysis of only individuals with European ancestry and a second meta-analysis of individuals with only East Asian ancestry ^36^ and found a concordant effect of lower *ACE* expression on increased schizophrenia risk in both datasets. The eQTL SNP used as an instrument for *ACE* expression (rs4277405) also has similar allele frequency across different ancestry populations (minor allele frequency of around 0.35 in Africans, Europeans, East and South Asians), and association in the GWAS is unlikely to be a result of uncorrected population stratification. The final assumption requires the genetic instrument to be associated with the outcome only through the exposure being tested. We have assessed association due to linkage or horizontal pleiotropy as well as assessed the likelihood of a common causal variant. **Figure 5** summarises the possible scenarios that may lead to the observed association between *ACE* expression and schizophrenia risk, and the analyses done to assess these scenarios.

**Figure 5.**
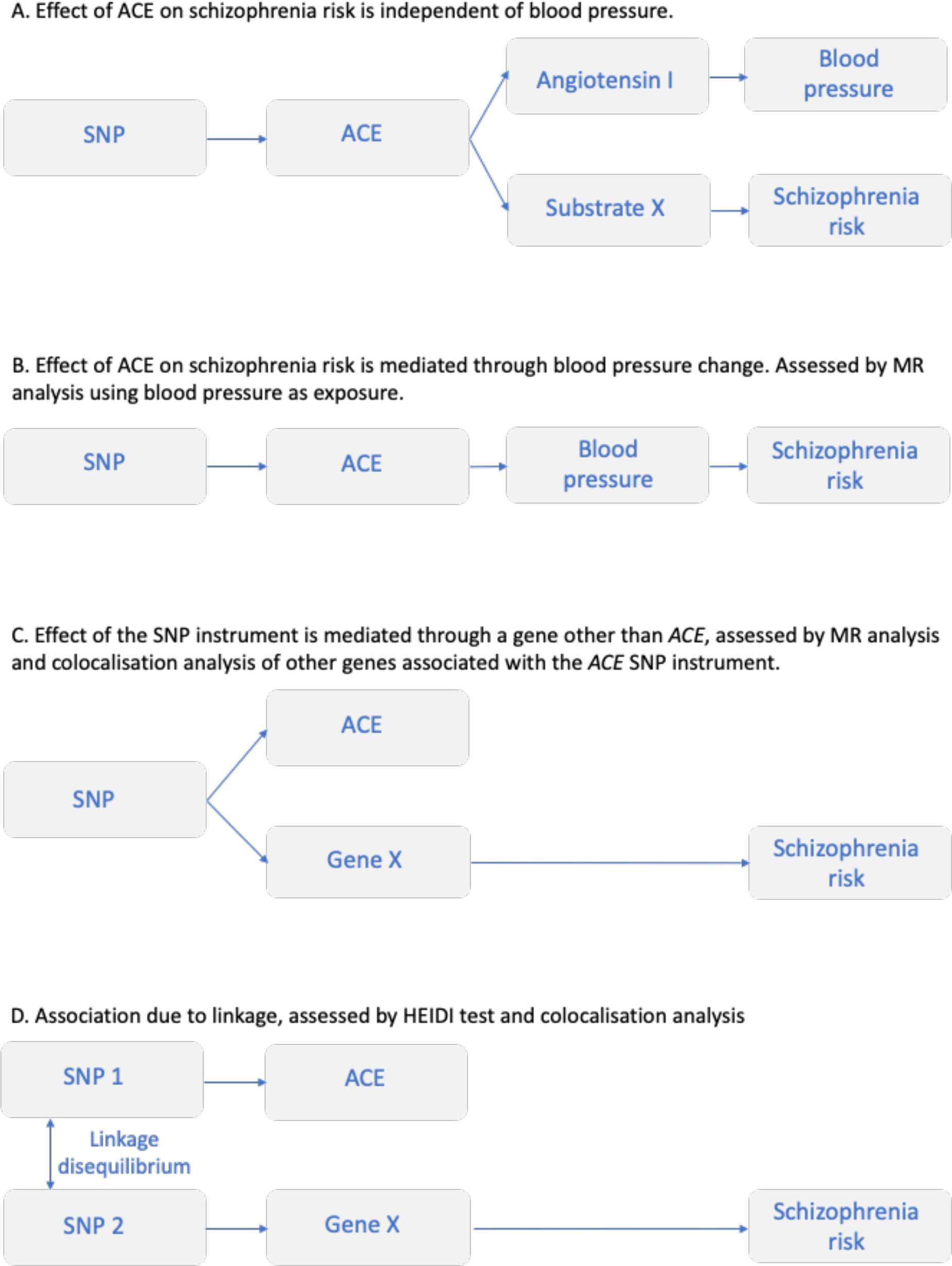
Possible explanations for observed association in MR analysis between *ACE* expression and schizophrenia risk.

Findings from MR analyses can provide prior information that could be used to improve the design of future studies. For example, most studies that have looked at antihypertensive drugs in relation to symptoms in psychiatric disorders have investigated drugs targeting the renin-angiotensin system (ARBs, ACE inhibitors and renin inhibitors) as a single group. Given that targeting different components of this pathway may have differential effects, future studies should look at individual drugs separately.

### Study limitations

MR analysis relies on the availability of a strong instrument for the exposure of interest. Using eQTL variants as instruments makes the assumption that changes in gene expression reflect changes in protein levels and/or protein activity, which may not always be the case. Therefore, lack of association in our analysis does not necessarily mean lack of biological effect of drug treatment, but rather lack of evidence that change in gene expression in the tested tissues is not associated with outcome. For *ACE*, we also use a second instrument that has been shown to be associated with both protein expression and activity in independent datasets, as well GWAS data on ACE protein levels in cerebral spinal fluid that show concordant effects of lower protein levels on increased risk. Though we have tried to assess horizontal pleiotropy, given the availability of only *cis* eQTL associations, we cannot assess any pleiotropy due to eQTL associations with genes located further away (*trans*-eQTLs). However, it is likely that any *trans-*effects are mediated through effects on the *cis*-gene^56,57^, in which case the MR assumption would still hold.

Tissue-specific effects of genes on disease can complicate interpretation of such analyses and restricting analysis to tissues that are most relevant to the disease of interest are more meaningful. In addition, different regions of the brain may have greater or lesser importance in disease aetiology. However, the power to conduct brain region-specific analysis is currently restricted by sample size of available eQTL datasets. Though relatively high correlation has been observed between blood and brain eQTLs^25^, any extrapolation will be restricted to genes that are expressed and have eQTLs in both. Though we found significant associations in cortex and cerebellum, given the small sample sizes, a null association could be attributed to lack of power, and the relevance of other brain regions cannot be ruled out.

Genetic variants reflect the effect of lifelong exposure on an outcome, therefore results from these analyses cannot be directly translated into effects of short-term drug treatment on disease risk or effects on neuropsychiatric symptoms in individuals that have already been diagnosed. On the other hand, this difference may prove useful, as it extends the duration of exposure well beyond what is feasible in randomised trials ^58^. RCTs to look at the effect of duration of drug treatment on disease incidence or on the positive and negative syndrome scale (PANSS) would be necessary to confirm such effects. It is important to note that disease risk here is based on a 1 SD change in *ACE* expression in blood, which was also associated with a 4.0 mmHg change in SBP. This change in SBP is comparable to the 4.7 mmHg reduction reported in a meta-analysis of RCTs for quarter-dose monotherapy compared to placebo^59^. However, different ACE inhibitors differ in lipophilicity and do not all cross the blood-brain barrier, which may be necessary for exerting effects on psychiatric disorders. Different ACE inhibitors also have been shown to have different affinities for the two functional domains of ACE^60^, making it difficult to extrapolate the actual effect of different ACE inhibitors from genetic analyses.

### Study implications

Current guidelines from the National Institute for Health Care and Excellence (NICE) recommend ACE inhibitors as first line treatment in individuals < 55 years of age or in individuals with type 2 diabetes (https://www.nice.org.uk/guidance/NG136). Disease onset for the majority of patients with schizophrenia usually occurs in late adolescence or early adult life, and such individuals are unlikely to have been treated for hypertension, ruling out the potential for medication as a cause in these individuals. However, if lower ACE activity plays a causal role in schizophrenia, then given the high prevalence of hypertension in these patients and therefore the need to treat with antihypertensive drugs, it would still be important to understand how ACE inhibitor exposure may affect symptoms, even if they were not cause. Any association between ACE inhibitors use and schizophrenia risk will mostly be relevant in late-onset (> 40 year of age), which accounts for around 20% of cases of schizophrenia^61^.

## Data Availability

The study has been conducted using publicly available data. Sources for each dataset are provided in Supplementary information.

## Acknowledgments

This study has been possible thanks to GWAS and eQTL summary statistics made publicly available by the Psychiatric Genetics Consortium (including GWAS summary statistics for major depression that include data from 23andMe), the eQTLGen Consortium, the GTEx Project, the PsychENCODE Project, Evangelou et al and Kauwe et al, as well as data from the UK Biobank Resource under Application Number 12505. SS is supported by a National Health and Medical Research Council (NHMRC) early career fellowship. SS, SC and NW are supported by NHMRC Program Grants 1078901 and 1113400.

## Contributions

SS conceived, designed, and supervised the overall project. SC performed analyses. SS and SC drafted the initial manuscript. All authors assisted with interpretation, commended on drafts of the manuscript, and approved the final version. SS attests that all listed authors meet authorship criteria and that no others meeting the criteria have been omitted.

